# Caspase-6-cleaved tau is relevant in Alzheimer’s disease but not in other tauopathies: diagnostic and therapeutic implications

**DOI:** 10.1101/2021.01.28.21250322

**Authors:** Panos Theofilas, Antonia M.H. Piergies, Song Hua Li, Cathrine Petersen, Alexander J. Ehrenberg, Rana A. Eser, Brian Chin, Teddy Yang, Shireen Khan, Raymond Ng, Salvatore Spina, Willian W. Seeley, Bruce L. Miller, Michelle R. Arkin, Lea T. Grinberg

**Affiliations:** Memory and Aging Center, UCSF Weill Institute for Neurosciences, University of California, San Francisco, San Francisco, CA, USA; Shanghai ChemPartner, Shanghai, China; ChemPartner San Francisco, South San Francisco, CA, USA; Department of Pathology, University of California, San Francisco, San Francisco, CA, USA; Global Brain Health Institute, University of California, San Francisco, San Francisco, CA, USA; Department of Pharmaceutical Chemistry and Small Molecule Discovery Center, UCSF, San Francisco, CA, USA; Department of Pathology, University of Sao Paulo Medical School, Sao Paulo, Brazil

**Keywords:** Alzheimer’s disease, tauopathies, tau cleavage, tau hyperphosphorylation, tau isoforms, caspase-6, cell counting, immunohistochemistry

## Abstract

**Aim:** Tau truncation (tr-tau) by active caspase-6 (aCasp-6) generates toxic tau fragments prone to self-aggregation. Yet, the relationship between aCasp-6, different forms of tr-tau, and hyperphosphorylated tau (p-tau) accumulation in human brains with Alzheimer’s disease (AD) and other tauopathies remains unclear.

**Methods:** We generated two neoepitope monoclonal antibodies against tr-tau sites (D402 and D13) targeted by aCasp-6. Then, we used 5-plex immunofluorescence to quantify the neuronal and astroglial burden of aCasp-6, tr-tau, p-tau, and their co-occurrence in healthy controls, AD, and primary tauopathies.

**Results:** Casp-6 activation was strongest in AD, followed by Pick’s disease (PiD), but almost absent in 4-repeat (4R) tauopathies. In neurons, the tr-tau burden was much more abundant in AD than in 4R tauopathies, and disproportionally higher when normalizing by p-tau pathology. Tr-tau astrogliopathy was detected in low numbers in 4R tauopathies. Unexpectedly, about half of tr-tau positive neurons in AD lacked p-tau aggregates.

**Conclusions:** Early modulation of aCasp-6 to reduce tr-tau pathology is a promising therapeutic strategy for AD, and possibly PiD, but is unlikely to benefit 4R tauopathies. The large percentage of tr-tau-positive neurons lacking p-tau suggests that not all neurons that are vulnerable to tau pathology are detected by a conventional p-tau Ser 202 antibody and that AD has distinct mechanisms of tangle formation. Therapeutic strategies against tr-tau pathology could be necessary to modulate tau abnormalities in AD. The disproportionally higher burden of tr-tau in AD supports the investigation of biofluid biomarkers against N-terminus tr-tau, which could detect AD and differentiate it from 4R tauopathies at a single patient level.

**3 - sentence summary:** Tau truncation (tr-tau) by active caspase-6 (aCasp-6) generates toxic tau fragments prone to self-aggregation, but the relationship between aCasp-6, tr-tau, and hyperphosphorylated tau (p-tau) accumulation in Alzheimer’s disease (AD) and other tauopathies remains unclear. We generated two neoepitope monoclonal antibodies against tr-tau sites (D402 and D13) targeted by aCasp-6 and used 5-plex immunofluorescence to quantify the neuronal and astroglial burden of aCasp-6, tr-tau, p-tau, and their co-occurrence in brains from healthy controls, AD, and primary tauopathies. We detected relatively high Casp-6 activation in AD, followed by Pick’s disease (PiD). aCasp-6 was almost absent in 4-repeat (4R) tauopathies, suggesting that early modulation of aCasp-6 to reduce tr-tau pathology is a promising therapeutic strategy in AD, and possibly PiD, but is unlikely to benefit 4R tauopathies.

## Introduction

Tau post-translational modifications (PTMs) define and modulate tau function in healthy and diseased states. Tauopathies, a major group of neurodegenerative diseases [1–3] including Alzheimer’s disease (AD), Pick’s disease (PiD), corticobasal degeneration (CBD), progressive supranuclear palsy (PSP), and argyrophilic grain disease (AGD), share a progressive accumulation of pathological tau in the brain. Although tauopathies are classically defined by the morphology and distribution of phosphorylated tau (p-tau) aggregates, other underexplored tau PTMs are also critical to tau pathogenesis [4–6].

Proteolytic truncation of tau (tr-tau) by active caspases has recently been recognized as a significant contributor to tau-driven nosology in AD and primary tauopathies [7–10]. Caspases (Casps), cysteine-aspartic proteases, are proteolytic enzymes with well-defined roles in apoptosis and inflammation [11, 12]. Pro-apoptotic caspase-6 (Casp-6) is an effector caspase, along with Casps-3 and -7. Most studies are directed at Casp-3 due to its central role in stress-induced apoptosis and neuronal death during development. However, previous studies, ours included, demonstrate relatively high levels of active caspase-6 (aCasp-6), but not -3, in the AD brain that co-occurs with an increase of abnormal proteins associated with the disease, such as the amyloid precursor protein and phospho-tau [13–16]. In AD, aCasp-6 acts downstream, promoting apoptosis [12, 17, 18], and upstream, by cleaving tau at confirmed and putative sites, like Asp421—a C-terminus site (D421) contributing to tau aggregation and the formation of tau neurofibrillary tangles (NFTs) [7, 9, 19].

We previously detected an increasing burden of neurons with aCasp-6 in the presence or absence of p-tau aggregates in AD starting at initial stages that paralleled the increasing burden of neurons with tau phosphorylated at Ser 202 [16]. This early and progressive increase of aCasp-6 levels in AD corroborates the hypothesis that modulation of Casp-6 activation and cleavage of tau may represent a promising therapeutic strategy in AD. However, fundamental gaps in our understanding of Casp-6 activation, tau cleavage, and the relationship between these processes and the formation of p-tau aggregates in neurons and astroglia persist. For example, it is unclear whether aCasp-6 is detected in tauopathies other than AD. Moreover, our knowledge on tr-tau burden in tauopathies and the extent to which tr-tau and p-tau aggregates co-occur in the same cells is practically limited to D421 in AD. D421 is not the only tr-tau form to accumulate in AD. Caspase-6 targets tau at other sites in AD, including D402 at the C-terminal and D13 at the N-terminal [14, 20, 21]. In AD, D402 levels in NFTs and neuropil threads were associated with lower global cognitive scores in cases with no history of cognitive impairment, suggesting that tau truncation may be an early event in AD pathogenesis [13]. Additionally, levels of D402 tr-tau (detected by polyclonal antibodies) correlate with pathological p-tau and neuronal loss in human brains affected by AD and has been tested as a CSF biomarker for AD since its levels positively correlate with AD severity [14, 20, 22]. *In vitro* studies have also shown that tau cleavage by aCasp-6 at D13, which is at the center of the Tau-12 and 5A6 epitopes, results in the loss of immunoreactivity with both Tau-12 and 5A6 N-terminal antibodies, suggesting a role for Casp-6 in the N-terminal truncation of tau [21]. However, the investigation of D13 and D402 tr-tau forms has been limited by the lack of reliable monoclonal antibodies (mAbs) targeting these specific cleavage sites.

To interrogate the spread of Casp-6 tr-tau in human tauopathies and the potential relevance of therapeutic modulation of Casp-6 activity in these diseases, we generated two novel epitope (neoepitope) mAbs targeting proteolytic sites predominantly cleaved by aCasp-6: D402 and D13. Next, we used multiplex immunofluorescence (IF) to quantitatively map neurons and astroglia with D402 and D13 tr-tau and evaluate their relationship to aCasp-6 and p-tau (Ser 202) aggregates in two cortical areas of a well-characterized postmortem brain cohort with AD, sporadic 3-repeat (3R), 4-repeat (4R) tauopathies, and healthy aging controls.

## Materials and Methods

### Participants

This study was exempt from ethical approval by the University of California, San Francisco (UCSF) institutional review board. Postmortem human brains were obtained from UCSF’s Neurodegenerative Disease Brain Bank [23]. All brains underwent standardized neuropathological assessment for neurodegenerative diseases that follow universally accepted guidelines [24, 25]. Inclusion criteria included a postmortem interval under 24 hours and a lack of more than one neuropathological diagnosis, an Axis I psychiatric disorder diagnosis, a non-dementia neurological disorder, and gross non-degenerative structural neuropathology. Control cases were free of any clinical symptoms of cognitive decline and neurological or non-incidental neuropathological diagnoses. To provide a broad picture of the most common sporadic tauopathies, our cohort of cases include 3R and 4R tauopathies–AD (3R/4R), AGD (4R), CBD (4R), PiD (3R), PSP (4R)–as well as clinical and pathological age-matched controls. Table 1 illustrates the characteristics of all 17 cases.

**Table 1.**
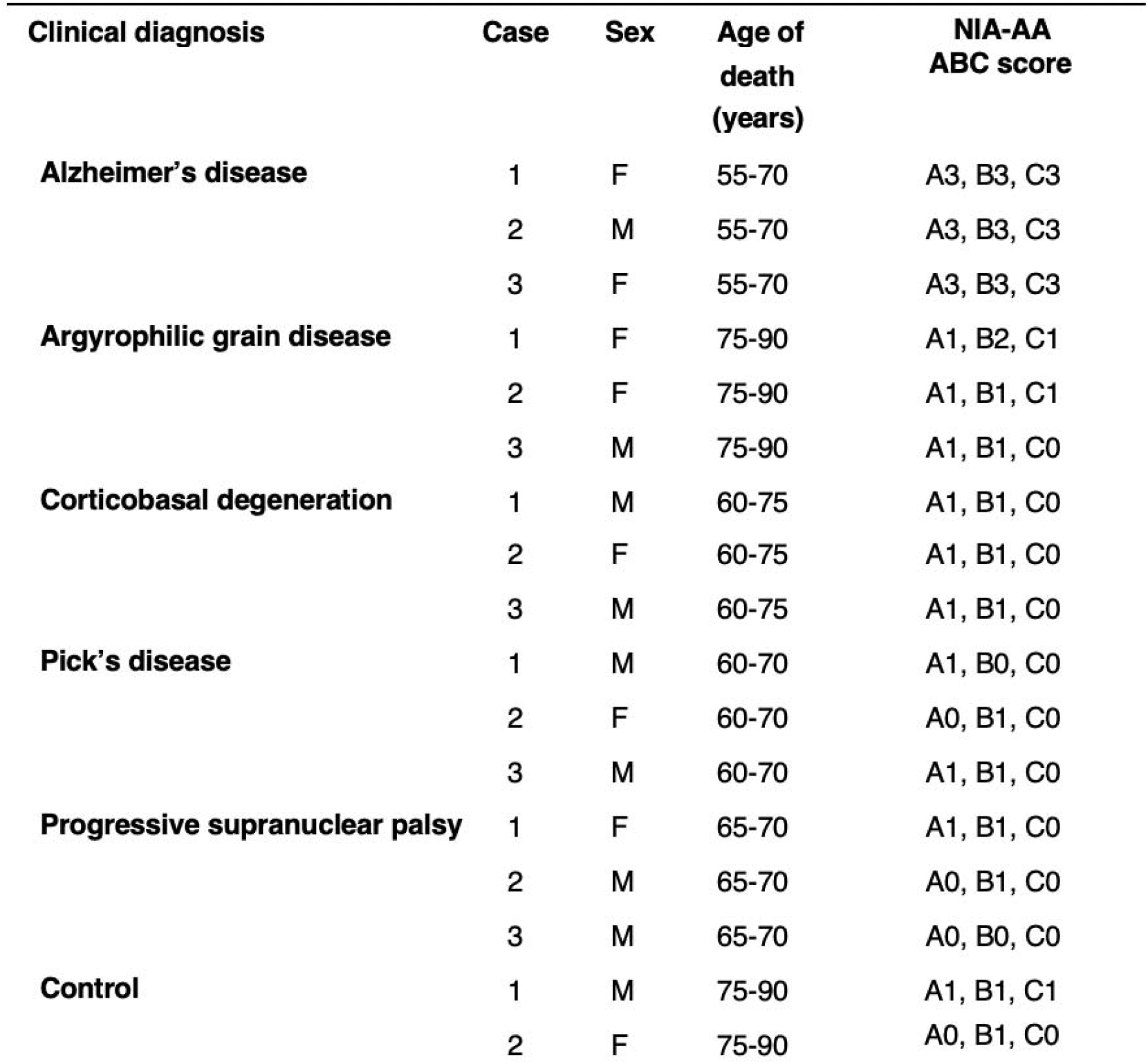
Demographic, clinical, and neuropathological characteristics of the 17 cases included in the study. Abbreviations: NIA-AA, National Institute on Aging and Alzheimer’s Association.

| Clinical diagnosis | Case | Sex | Age of death (years) | NIA-AA ABC score |
| --- | --- | --- | --- | --- |
| **Alzheimer's disease** | 1 | F | 55-70 | A3, B3, C3 |
|  | 2 | M | 55-70 | A3, B3, C3 |
|  | 3 | F | 55-70 | A3, B3, C3 |
| **Argyrophilic grain disease** | 1 | F | 75-90 | A1, B2, C1 |
|  | 2 | F | 75-90 | A1, B1, C1 |
|  | 3 | M | 75-90 | A1, B1, C0 |
| **Corticobasal degeneration** | 1 | M | 60-75 | A1, B1, C0 |
|  | 2 | F | 60-75 | A1, B1, C0 |
|  | 3 | M | 60-75 | A1, B1, C0 |
| **Pick's disease** | 1 | M | 60-70 | A1, B0, C0 |
|  | 2 | F | 60-70 | A0, B1, C0 |
|  | 3 | M | 60-70 | A1, B1, C0 |
| **Progressive supranuclear palsy** | 1 | F | 65-70 | A1, B1, C0 |
|  | 2 | M | 65-70 | A0, B1, C0 |
|  | 3 | M | 65-70 | A0, B0, C0 |
| **Control** | 1 | M | 75-90 | A1, B1, C1 |
|  | 2 | F | 75-90 | A0, B1, C0 |

### Development of caspase-6-cleaved tau neoepitope monoclonal antibodies

Except for TauC3 (D421), mAbs against Casp-6 tr-tau are not available. In collaboration with ChemPartner, we generated two neoepitope mAbs targeting tau cleaved by Casp-6: mAbD13 (14-442; Peptide I: HAGTYGLGDRKC) and mAbD402 (1-401; Peptide II: CIVYKSPVVSGD) as previously described [26]. Briefly, both mAbs were produced by immunizing 6-to 8-week-old wild type Balb/c, and SJL mice (SLAC) with keyhole limpet hemocyanin (KLH) conjugated tau peptides using protocols approved by the ChemPartner IACUC committee. 50 µg of each peptide was injected into each mouse’s abdominal cavity along with 0.25 mL of Complete Freund’s Adjuvant (Sigma). Blood samples were collected one week after immunization. The antibody titer and specificity in serum were determined by enzyme-linked immunosorbent assay (ELISA) analysis against BSA-conjugated peptides I and II and western blot against full-length tau, tau 1-402, and tau 14-441 recombinant proteins. Mice with specific immune responses against tau peptides were selected for fusion and were given a final boost by intraperitoneal injection of 100 µg of the corresponding immunogen. After four days, all mice were sacrificed, and their splenocytes lysed in NH4OH at 1% (w/w), followed by centrifugation at 1000 rpm and washes with DMEM (Invitrogen). Viable splenocytes were fused with mouse myeloma cells SP2/0 (ATCC) at a ratio of 5:1 with high-efficiency electric fusion (BTX ECM200). Fused cells were re-suspended in DMEM with 20% FBS and hypoxanthine-aminopterin-thymidine (HAT) medium (Invitrogen). 14 days after cell fusion, hybridoma supernatants were collected and screened by ELISA. Clones with an OD450 nm > 1.0 were expanded in a 24-well plate containing DMEM with 10% heat-inactivated FBS, and supernatants were collected after three days of culture. The antibody isotypes were determined, and ELISA and western blot were used to test their ability to bind to tau. Clones that showed desired reactivity and specificity against tau were subjected to subcloning to get stable monoclonal hybridoma cells. Sub-cloning was carried out by limited dilution in a 96-well plate with DMEM media containing 10% FBS. Clones with specific tau binding were further expanded in DMEM media containing 10% FBS for subsequent antibody production and cryopreservation. See Supplemental Experimental Procedures for the analysis of antibody specificity.

### Tissue processing and multiplex immunofluorescence staining

Immunofluorescence (IF) assays are prone to batch variations. To minimize this risk, we created tissue microarrays (TMAs) by using a 5 mm biopsy punch to sample the middle frontal gyrus (MFG) at the level of the optical chiasm and the inferior temporal gyrus (ITG) at the level of anterior commissure from all 17 cases. The longest punch axis was always perpendicular to the gyrus surface to avoid oblique sampling. Each TMA accommodated six cases that were embedded in a single paraffin block. Eight μm thick sections of each TMA were mounted on histological slides before undergoing multiplex IF, through which we would detect neurons (NeuN), p-tau (CP13), aCasp-6, and Casp-6-tr-tau (see Table S1 for a list of primary antibodies). We chose CP13 as a proxy for p-tau because the phosphorylation of tau at Ser 202 occurs earlier in AD, is present in all tauopathies, and rarely occurs in healthy brains [27–30]. All IF assays were performed using a Ventana Discovery Ultra automated staining instrument (Ventana Medical Systems). The general procedure and quality control steps of our IF protocol have been described in detail elsewhere [31]. Briefly, tissue sections were baked at 60°C for 20 minutes and subsequently deparaffinized via incubation in 69°C Ventana’s Discovery wash solution for 24 minutes. Epitope retrieval was accomplished by incubating sections in a 95°C Discovery CC1 solution (Ventana) for 56 minutes. Following epitope retrieval, sections were incubated in room temperature Discovery inhibitor (Ventana) for 16 minutes to inactivate endogenous horseradish peroxidase (HRP). See Supplemental Experimental Procedures for antibody labeling approach.

### Quantitative analysis of positive markers

Cell quantification was performed blinded to clinical and neuropathological diagnosis and reviewed by three of the authors independently. Each tissue punch was imaged at 20x magnification with a Zeiss AxioImager A2 microscope equipped with a Zeiss Colibri 7: Type FRR [G/Y] CBV-UC 7-channel fluorescence light source and an electronic platform. AF350 was visualized using a DAPI filter set, AF488 with a GFP filter set, AF546 with a DsRed filter set, AF647 with a Cy5 filter set, and AF790 with a Cy7 filter set. Figures 1 and 2 show representative examples of positive markers in neurons (all tauopathies) and astroglia (CBD and PSP), respectively. A semi-automated pipeline was used to quantify marker positivity in both cell types (Figure 3). This approach is further described in Supplemental Experimental Procedures.

**Figure 1.**
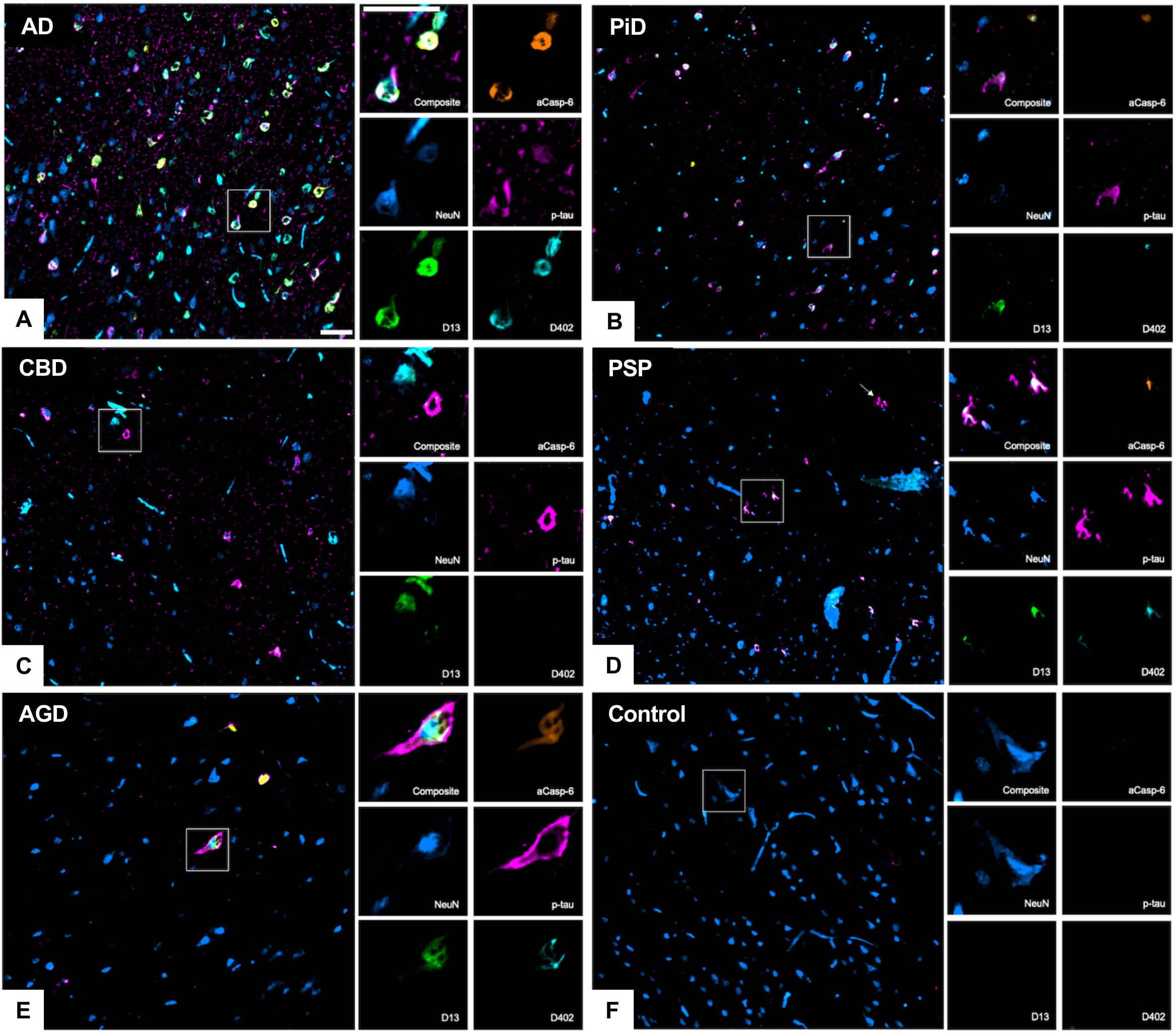
Neuronal marker positivity in the middle frontal and inferior temporal gyrus of human postmortem brains with common tauopathies. Low and high magnification images showing multiplex immunofluorescence staining of neurons (NeuN, blue), active caspase-6 (aCasp-6, orange), neoepitope mAbs of caspase-6-cleaved tau sites (D402 and D13, green and cyan, respectively), and phosphorylated tau (Ser 202; p-tau, magenta). Compared to other tauopathies (B-E), AD (A) showed the most robust positivity for all markers, while no or negligible positivity was observed in controls (F). Abbreviations: AD, Alzheimer’s disease (A); PiD, Pick’s disease (B); CBD, corticobasal degeneration (C); PSP progressive supranuclear palsy (D); AGD, argyrophilic grain disease. Scale bars: 50 μm.

**Figure 2.**
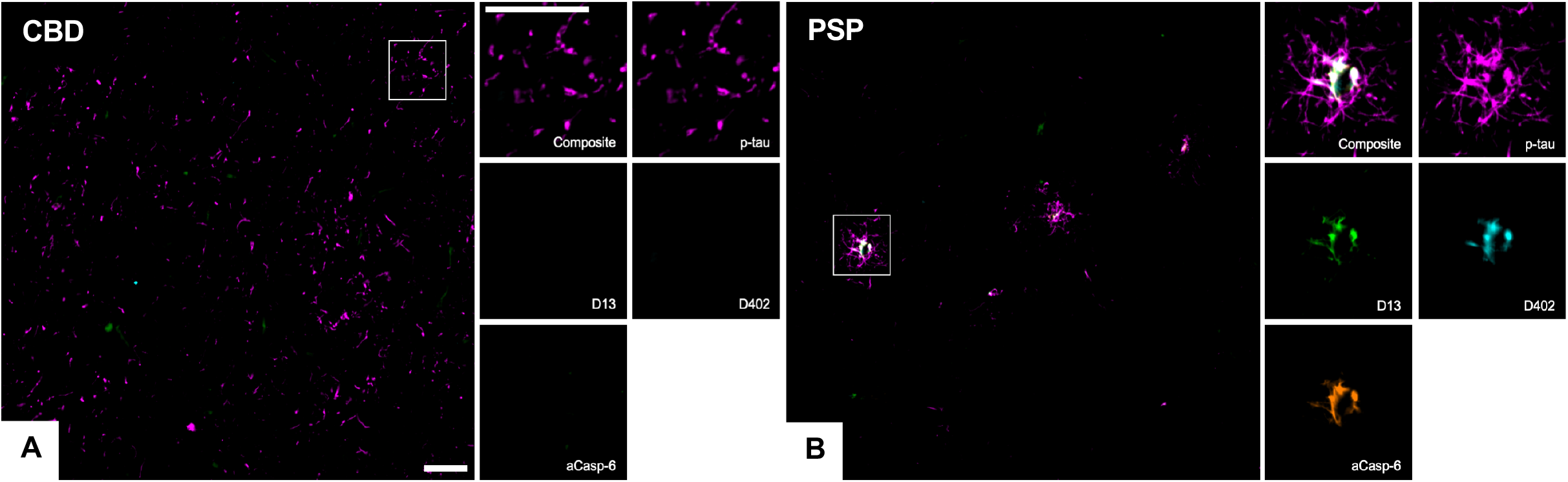
Multiplex immunofluorescence images showing the co-occurrence of phosphorylated-tau (Ser202, magenta), active caspase-6 (aCasp-6, orange), and neoepitope mAbs against caspase-6-cleaved tau sites (D402 and D13, green and cyan, respectively) in astroglial cells from human postmortem brains affected by CBD (A) and PSP (B). PSP (B) showed greater positivity for all markers, as compared to CBD (A). Abbreviations: CBD, corticobasal degeneration (A); PSP progressive supranuclear palsy (B). Scale bars: 50 μm.

**Figure 3.**
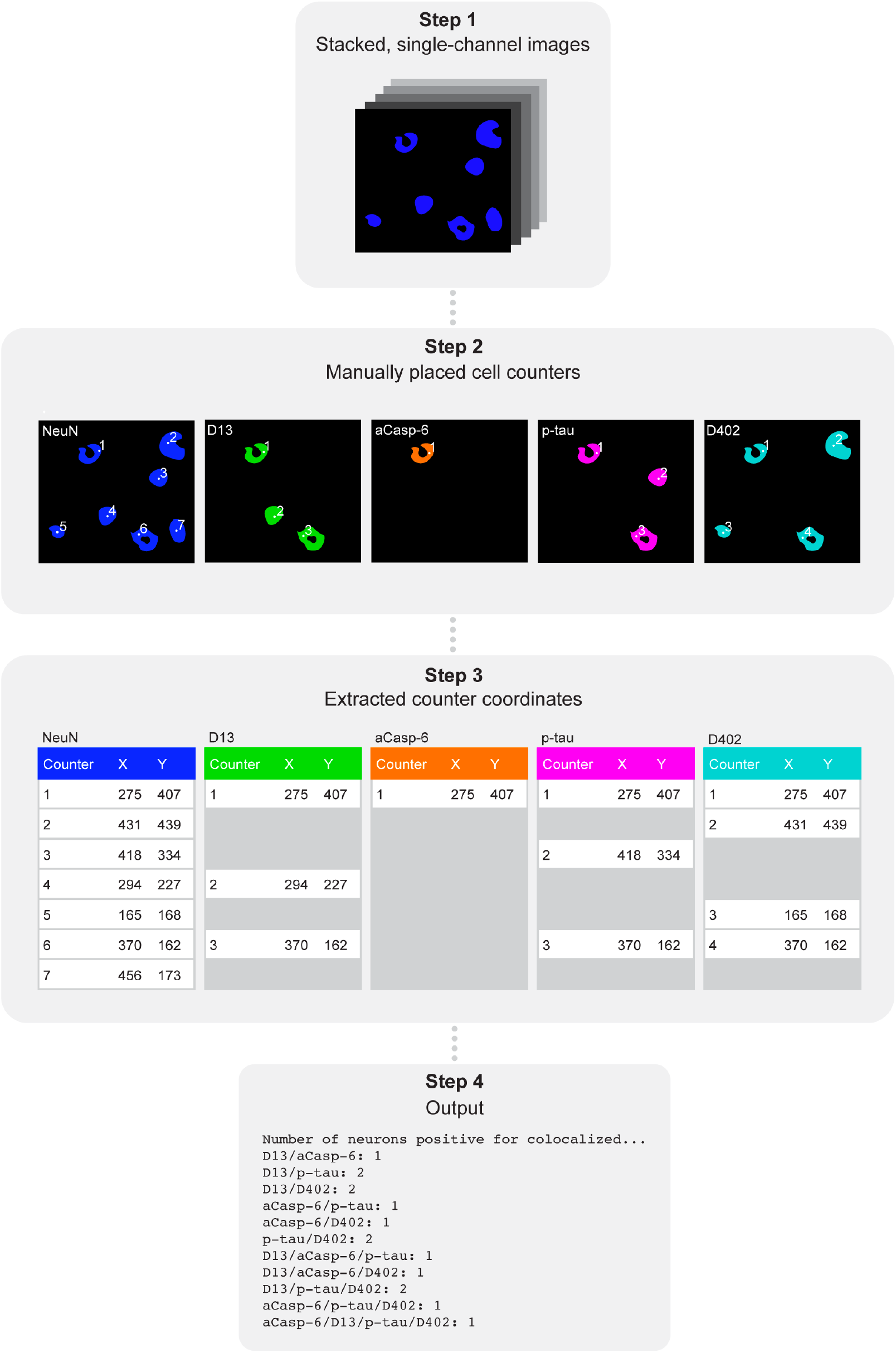
Schematic of the semi-automated cell counting approach. First, single-channel images were stacked (step one) followed by manual placement of counters on positive cells using FIJI’s built-in counting tool (step two). Next, counter coordinate data was extracted from FIJI files (step three). The number and type of co-occurring markers were determined by searching for coordinates present within a two-pixel radius of each other using a Python script (step four). Abbreviations: aCasp-6, active caspase-6; p-tau, phosphorylated tau.

### Statistical analysis

Data analysis was performed using R Statistical Software (version 3.6.1; R Foundation for Statistical Computing, Vienna, Austria). Means and standard deviations were calculated for demographic and quantitative neuropathological data.

## Results

Table 1 summarizes the demographic, clinical, and neuropathological characteristics of all 17 cases used in this study. 52.94% were male, the mean (SD) age of death was 70.82 (9.29) years, the mean (SD) postmortem interval was 10.18 (3.96) hours, and the mean (SD) brain weight was 1148.94 (124.79) grams.

### All tauopathies show expected positivity for neuronal p-tau

We used the p-tau mAb CP13 (Table S1) to map p-tau Ser 202-positive neurons in five sporadic tauopathies and healthy controls. All cases showed expected percentages of neuronal p-tau inclusions in the MFG and ITG. In the MFG, the absence of neuronal p-tau in AGD was anticipated since this region is typically spared of tau pathology. Of note, in controls, we found p-tau positivity in an average of 0.09% of ITG neurons, which is within the bounds expected of non-pathological aging. Overall, our results confirm the presence of tau lesions in predicted regions based on disease diagnosis and provide a basis for us to interpret the magnitude of tr-tau burden (Table 2; Figures 4, 5).

**Table 2.**
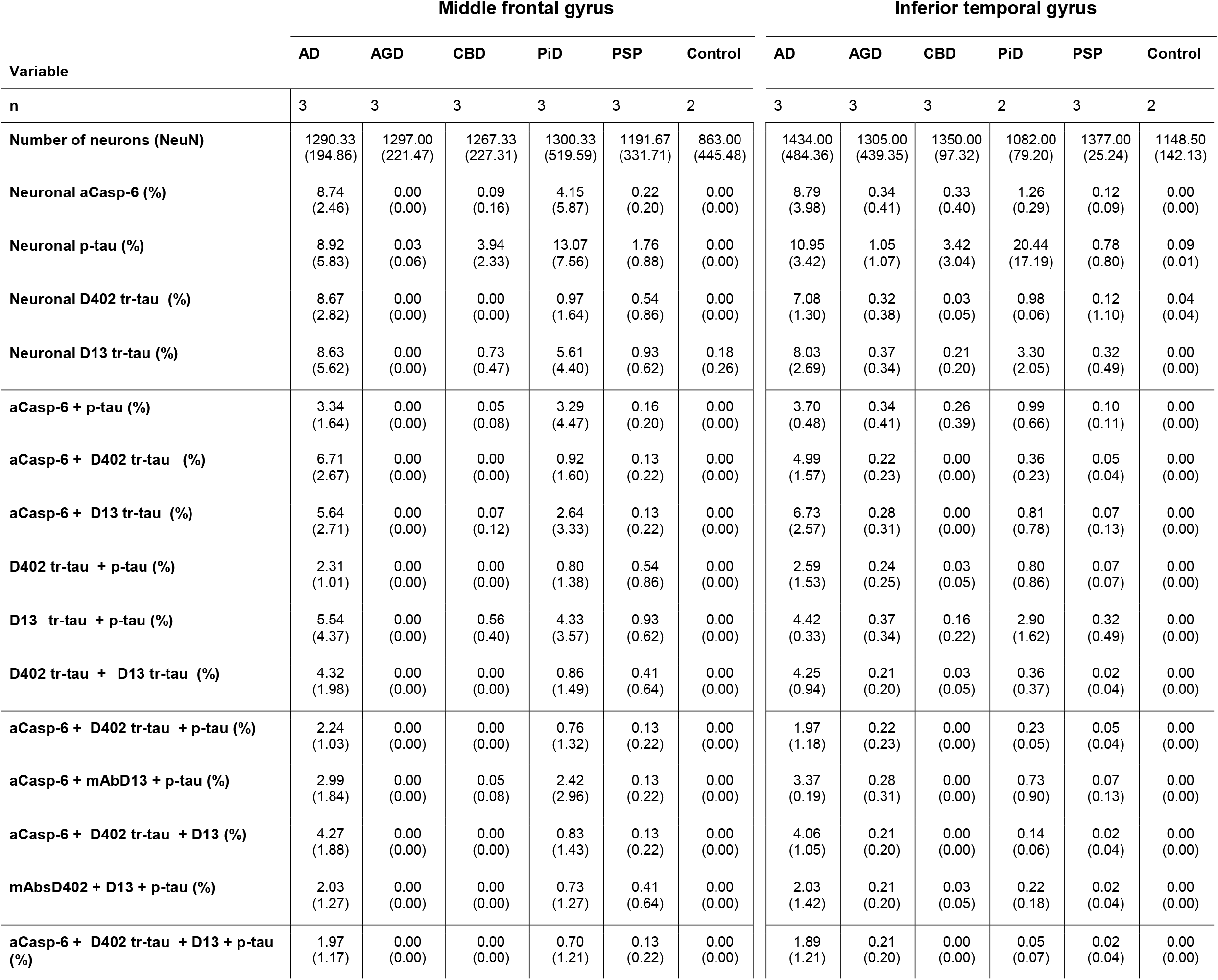
Mean (SD) percentage of neurons positive for the active caspase-6, D402 truncated tau, D13 truncated tau, phosphorylated tau, individually and in combination. All cases showed expected neuronal p-tau positivity, based on their diagnosis. AD cases showed strongest positivity for neuronal caspase-6, followed by PiD. AD and PiD cases showed the highest percentage of neurons positive for mAbsD402 and D13. Numbers for individual markers include neurons that are positive for more than one marker. Abbreviations: aCasp-6, active caspase-6; AD, Alzheimer’s disease; AGD, argyrophilic grain disease; CBD, corticobasal degeneration; PiD, Pick’s disease; PSP, progressive supranuclear palsy; p-tau, phosphorylated tau.

| Variable | Middle frontal gyrus |  |  |  |  |  | Inferior temporal gyrus |  |  |  |  |  |
| --- | --- | --- | --- | --- | --- | --- | --- | --- | --- | --- | --- | --- |
|  | AD | AGD | CBD | PiD | PSP | Control | AD | AGD | CBD | PiD | PSP | Control |
| n | 3 | 3 | 3 | 3 | 3 | 2 | 3 | 3 | 3 | 2 | 3 | 2 |
| **Number of neurons (NeuN)** | 1290.33<br>(194.86) | 1297.00<br>(221.47) | 1267.33<br>(227.31) | 1300.33<br>(519.59) | 1191.67<br>(331.71) | 863.00<br>(445.48) | 1434.00<br>(484.36) | 1305.00<br>(439.35) | 1350.00<br>(97.32) | 1082.00<br>(79.20) | 1377.00<br>(25.24) | 1148.50<br>(142.13) |
| **Neuronal aCasp-6 (%)** | 8.74<br>(2.46) | 0.00<br>(0.00) | 0.09<br>(0.16) | 4.15<br>(5.87) | 0.22<br>(0.20) | 0.00<br>(0.00) | 8.79<br>(3.98) | 0.34<br>(0.41) | 0.33<br>(0.40) | 1.26<br>(0.29) | 0.12<br>(0.09) | 0.00<br>(0.00) |
| **Neuronal p-tau (%)** | 8.92<br>(5.83) | 0.03<br>(0.06) | 3.94<br>(2.33) | 13.07<br>(7.56) | 1.76<br>(0.88) | 0.00<br>(0.00) | 10.95<br>(3.42) | 1.05<br>(1.07) | 3.42<br>(3.04) | 20.44<br>(17.19) | 0.78<br>(0.80) | 0.09<br>(0.01) |
| **Neuronal D402 tr-tau (%)** | 8.67<br>(2.82) | 0.00<br>(0.00) | 0.00<br>(0.00) | 0.97<br>(1.64) | 0.54<br>(0.86) | 0.00<br>(0.00) | 7.08<br>(1.30) | 0.32<br>(0.38) | 0.03<br>(0.05) | 0.98<br>(0.06) | 0.12<br>(1.10) | 0.04<br>(0.04) |
| **Neuronal D13 tr-tau (%)** | 8.63<br>(5.62) | 0.00<br>(0.00) | 0.73<br>(0.47) | 5.61<br>(4.40) | 0.93<br>(0.62) | 0.18<br>(0.26) | 8.03<br>(2.69) | 0.37<br>(0.34) | 0.21<br>(0.20) | 3.30<br>(2.05) | 0.32<br>(0.49) | 0.00<br>(0.00) |
| **aCasp-6 + p-tau (%)** | 3.34<br>(1.64) | 0.00<br>(0.00) | 0.05<br>(0.08) | 3.29<br>(4.47) | 0.16<br>(0.20) | 0.00<br>(0.00) | 3.70<br>(0.48) | 0.34<br>(0.41) | 0.26<br>(0.39) | 0.99<br>(0.66) | 0.10<br>(0.11) | 0.00<br>(0.00) |
| **aCasp-6 + D402 tr-tau (%)** | 6.71<br>(2.67) | 0.00<br>(0.00) | 0.00<br>(0.00) | 0.92<br>(1.60) | 0.13<br>(0.22) | 0.00<br>(0.00) | 4.99<br>(1.57) | 0.22<br>(0.23) | 0.00<br>(0.00) | 0.36<br>(0.23) | 0.05<br>(0.04) | 0.00<br>(0.00) |
| **aCasp-6 + D13 tr-tau (%)** | 5.64<br>(2.71) | 0.00<br>(0.00) | 0.07<br>(0.12) | 2.64<br>(3.33) | 0.13<br>(0.22) | 0.00<br>(0.00) | 6.73<br>(2.57) | 0.28<br>(0.31) | 0.00<br>(0.00) | 0.81<br>(0.78) | 0.07<br>(0.13) | 0.00<br>(0.00) |
| **D402 tr-tau + p-tau (%)** | 2.31<br>(1.01) | 0.00<br>(0.00) | 0.00<br>(0.00) | 0.80<br>(1.38) | 0.54<br>(0.86) | 0.00<br>(0.00) | 2.59<br>(1.53) | 0.24<br>(0.25) | 0.03<br>(0.05) | 0.80<br>(0.86) | 0.07<br>(0.07) | 0.00<br>(0.00) |
| **D13 tr-tau + p-tau (%)** | 5.54<br>(4.37) | 0.00<br>(0.00) | 0.56<br>(0.40) | 4.33<br>(3.57) | 0.93<br>(0.62) | 0.00<br>(0.00) | 4.42<br>(0.33) | 0.37<br>(0.34) | 0.16<br>(0.22) | 2.90<br>(1.62) | 0.32<br>(0.49) | 0.00<br>(0.00) |
| **D402 tr-tau + D13 tr-tau (%)** | 4.32<br>(1.98) | 0.00<br>(0.00) | 0.00<br>(0.00) | 0.86<br>(1.49) | 0.41<br>(0.64) | 0.00<br>(0.00) | 4.25<br>(0.94) | 0.21<br>(0.20) | 0.03<br>(0.05) | 0.36<br>(0.37) | 0.02<br>(0.04) | 0.00<br>(0.00) |
| **aCasp-6 + D402 tr-tau + p-tau (%)** | 2.24<br>(1.03) | 0.00<br>(0.00) | 0.00<br>(0.00) | 0.76<br>(1.32) | 0.13<br>(0.22) | 0.00<br>(0.00) | 1.97<br>(1.18) | 0.22<br>(0.23) | 0.00<br>(0.00) | 0.23<br>(0.05) | 0.05<br>(0.04) | 0.00<br>(0.00) |
| **aCasp-6 + mAbD13 + p-tau (%)** | 2.99<br>(1.84) | 0.00<br>(0.00) | 0.05<br>(0.08) | 2.42<br>(2.96) | 0.13<br>(0.22) | 0.00<br>(0.00) | 3.37<br>(0.19) | 0.28<br>(0.31) | 0.00<br>(0.00) | 0.73<br>(0.90) | 0.07<br>(0.13) | 0.00<br>(0.00) |
| **aCasp-6 + D402 tr-tau + D13 (%)** | 4.27<br>(1.88) | 0.00<br>(0.00) | 0.00<br>(0.00) | 0.83<br>(1.43) | 0.13<br>(0.22) | 0.00<br>(0.00) | 4.06<br>(1.05) | 0.21<br>(0.20) | 0.00<br>(0.00) | 0.14<br>(0.06) | 0.02<br>(0.04) | 0.00<br>(0.00) |
| **mAbsD402 + D13 + p-tau (%)** | 2.03<br>(1.27) | 0.00<br>(0.00) | 0.00<br>(0.00) | 0.73<br>(1.27) | 0.41<br>(0.64) | 0.00<br>(0.00) | 2.03<br>(1.42) | 0.21<br>(0.20) | 0.03<br>(0.05) | 0.22<br>(0.18) | 0.02<br>(0.04) | 0.00<br>(0.00) |
| **aCasp-6 + D402 tr-tau + D13 + p-tau (%)** | 1.97<br>(1.17) | 0.00<br>(0.00) | 0.00<br>(0.00) | 0.70<br>(1.21) | 0.13<br>(0.22) | 0.00<br>(0.00) | 1.89<br>(1.21) | 0.21<br>(0.20) | 0.00<br>(0.00) | 0.05<br>(0.07) | 0.02<br>(0.04) | 0.00<br>(0.00) |

**Figure 4.**
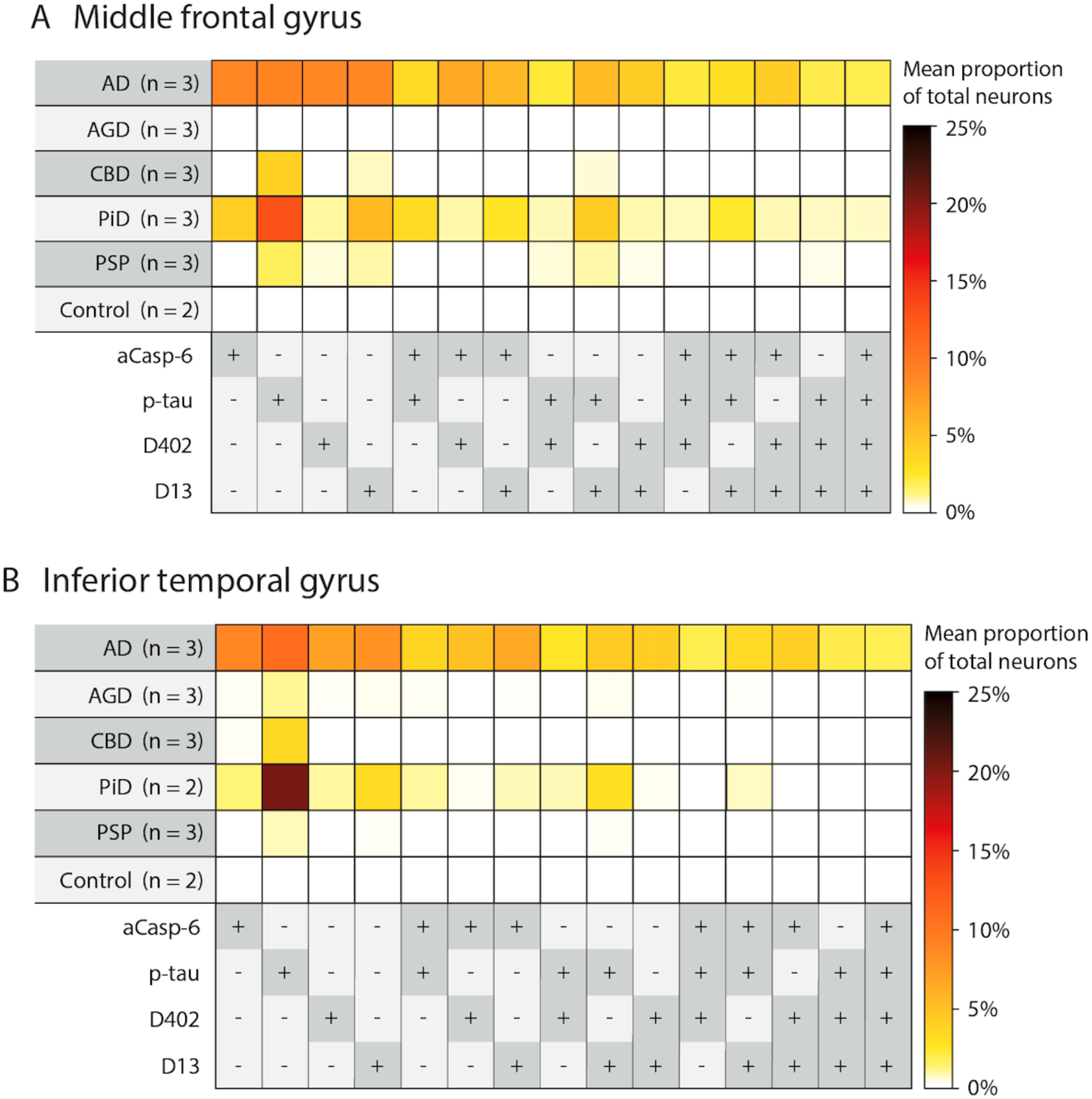
Heatmaps illustrating mean percentages of neurons in the middle frontal (A) and inferior temporal gyrus (B) with antibody positivity for active caspase-6 (aCasp-6), mAbD402, mAbD13, and phosphorylated tau (Ser202; p-tau). AD, AGD, CBD, PiD, and PSP cases showed expected p-tau positivity in neurons, based on diagnosis. AD brains exhibited the highest neuronal positivity for aCasp-6, mAbD402, and mAbD13, while PiD showed relatively moderate positivity for these markers. AGD showed no positivity for aCasp-6, mAbD402, or mAbD13 in the middle frontal gyrus and negligible positivity in the inferior temporal gyrus. CBD and PSP showed minimal positivity for aCasp-6, D402, and D13. Positivity for aCasp-6, mAb402, and mAbD13 was either not detected or scarce in controls. Abbreviations: AD, Alzheimer’s disease; AGD, argyrophilic grain disease; CBD, corticobasal degeneration; PiD, Pick’s disease; PSP, progressive supranuclear palsy. Rows represent tauopathies, controls, and a list of antibodies included in the analysis. Cells represent mean percentages of total neurons (color gradient) found positive for individual or multiple antibodies. The +/- symbols represent the presence (+, dark grey) or absence (-, light grey) of antibody signal.

**Figure 5.**
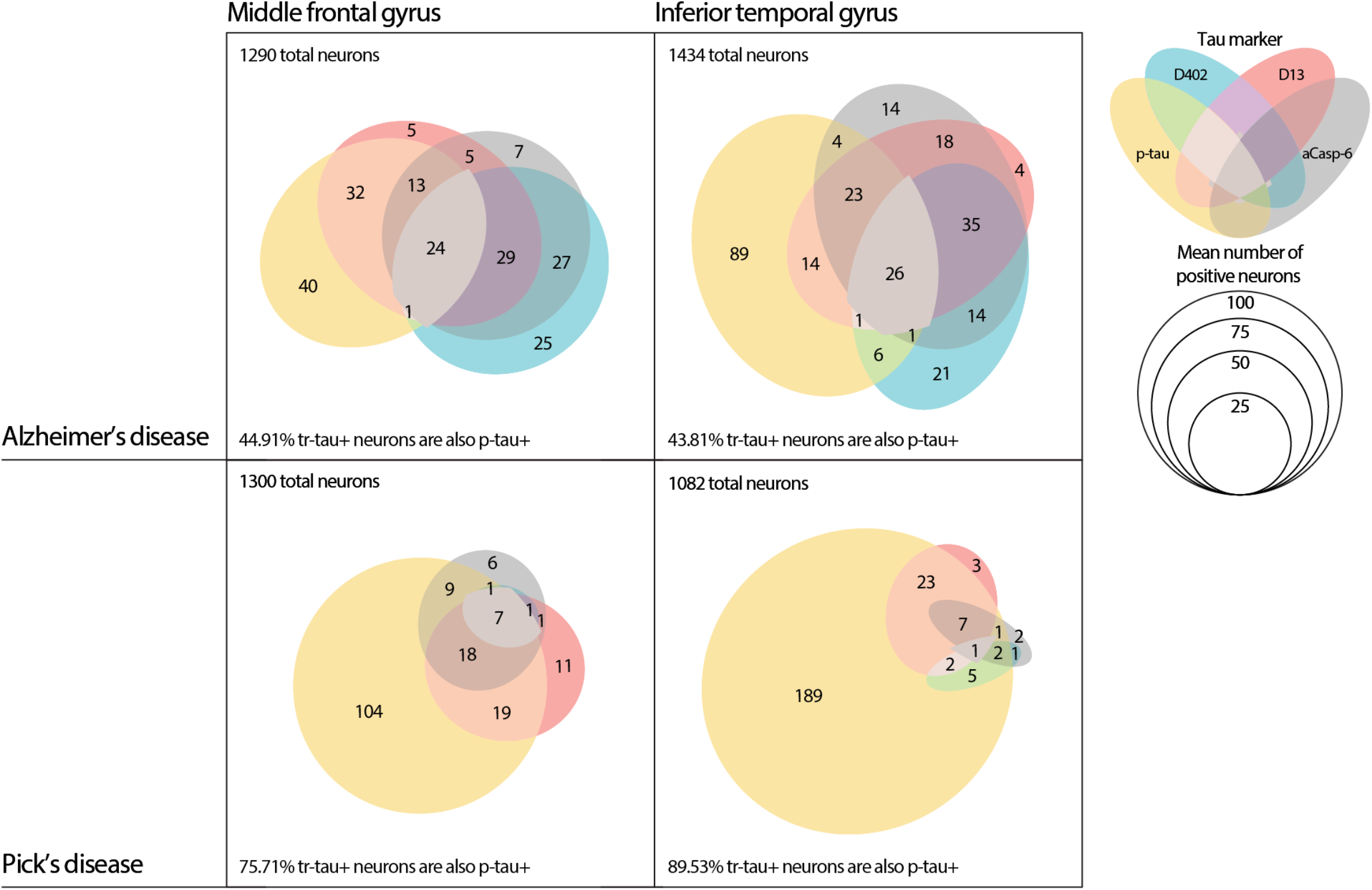
Venn diagrams illustrating the frequency of marker co-occurrence for AD and PiD, the two tauopathies with the highest marker positivity. Each colored ellipse represents mean numbers of positive neurons for each antibody, including phosphorylated tau (yellow), mAbs D402 (cyan) and D13 (orange), and active caspase-6 (grey) identified by multiplex immunofluorescence. Abbreviations: AD, Alzheimer’s disease; PiD, Pick’s disease; aCasp-6; active caspase-6; p-tau, phosphorylated tau.

### In AD, the percentage of neurons positive for aCasp-6 is at least 2.11 times higher than in all other tauopathies tested

In AD, aCasp-6 promotes tau cleavage. We previously found that the percentage of neurons with aCasp-6 increases as the disease progresses [16]. Here, we asked if this increase is a feature of other common tauopathies (Table 2; Figures 4, 5). In AD, we found that an average of 8.74% neurons in the MFG and 8.79% neurons in ITG were positive for aCasp-6. In control cases, we did not find any aCasp-6 positivity in either region. The percentages of MFG and ITG neurons with aCasp-6 positivity was much higher in AD than in all other tauopathies tested. PiD, the tauopathy with the second highest percentage of neurons with aCasp-6 positivity, only had aCasp-6 in 4.15% of MFG neurons and 1.26% of ITG neurons. These findings demonstrate that neuronal caspase-6 activity seems to be strongest in AD, relatively moderate in PiD, and negligible in other tauopathies, pointing to potential differences in Casp-dependent tau cleavage and, therefore, therapeutic approaches in AD and non-AD tau.

### Neuronal Casp-6-cleaved tau aggregates are more prominent in AD and PiD than in 4-repeat tauopathies

Previous studies focusing on the tr-tau sites D421 (mAbTauC3) and D402 (polyclonal antibody) showed tr-tau aggregates in AD, but relatively little is known about tr-tau aggregates in common primary tauopathies. Here, we used neoepitope mAbs targeting tau cleaved by Casp-6 at a C-terminus site (D402) and a previously unexplored N-terminus site (D13) to quantify the magnitude of tr-tau pathology in AD and other tauopathies (Figures 4, 5). Overall, AD and PiD exhibited at least 1.80x greater neuronal positivity for mAbD402 than the other tauopathies, reaching as much as 8.67% in MFG neurons and 7.08% ITG neurons in AD. In AD, the percentages were similar for mAbD13 (8.63% of MFG neurons and 8.03% of ITG neurons). Although the percentages of D13 tr-tau neurons were also noteworthy in PiD—with positivity in 5.61% of MFG neurons and 3.30% of ITG neurons—D402 tr-tau was less abundant (present in only 0.97% of MFG neurons and 0.98% of ITG neurons). All other diseases showed minimal percentages (<1%) of mAbD402 or mAbD13 positivity in neurons (Table 2; Figures 4, 5). These results suggest that Casp-6-tr-tau is a more defining feature of tauopathies with 3R tau aggregates (AD and PiD) than 4R tau aggregates (AGD, CBD, and PSP). Also, it highlights the potential of measuring D13 tr-tau levels as a potential fluid-based biomarker to differentiate AD from other tauopathies.

### In AD, the tauopathy with the highest percentage of aCasp-6-positive neurons, 89.6% of these neurons also harbor tr-tau

Co-occurrence was lower in the opposite direction, as only 72.70-76.45% (mAbD402) and 65.44-84.38% (mAbD13) of tr-tau neurons were also positive for aCasp-6 in AD. Intriguingly, in PiD, only about 1/3 of tr-tau positive neurons were also positive for aCasp-6 (Table 2; Figures 4, 5).

### In AD, a substantial percentage of neurons with tr-tau lack p-tau (Ser 202) inclusions

P-tau inclusions are a hallmark of all tauopathies, and abnormal tau phosphorylation at Ser202 is considered one of the earliest and most universal events in tau pathogenesis. Here, we evaluated the degree of co-occurrence between p-tau and tr-tau at D402 and D13 cleavage sites (Table 2; Figures 4, 5). Since the percentage of neurons with D402 and D13 tr-tau were minimal in the tested 4R tauopathies (AGD, CBD, PSP), we focused on AD and PiD. Regarding the co-occurrence of tr-tau and p-tau, in PiD, 80.65% and 76.30% of MFG neurons positive for D402 and D13 tr-tau, respectively, also showed p-tau positivity. Percentages were similar in ITG neurons (81.82% and 87.67%). Surprisingly, in AD, only 32.81% and 52.59% of MFG neurons and 26.30% and 64.22% of ITG neurons positive for D402 and D13 tr-tau showed p-tau positivity. These results were unexpected and suggest that a substantial population of neurons undergoes abnormal tau changes that would not be captured in studies only focusing on p-tau. Furthermore, they reveal a neglected area of investigation related to selective vulnerability and pathogenesis of AD and, to a lesser extent, PiD. Interestingly, D402 and D13 tr-tau only show partial neuronal co-occurrence in AD (Table 2; Figures 4, 6).

**Figure 6.**
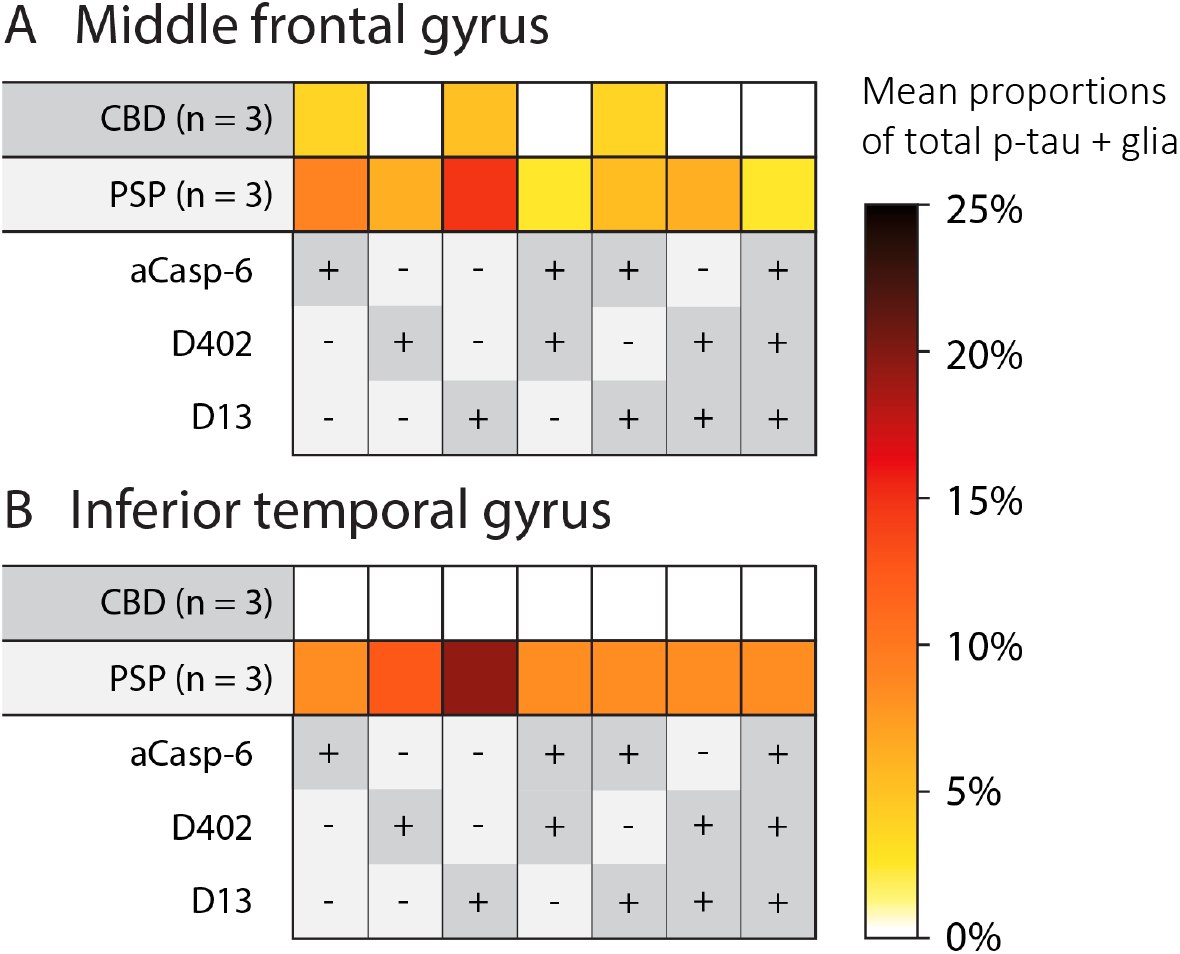
Heatmaps illustrating mean percentages of phospho-tau positive astroglia, in the middle frontal (A) and inferior temporal gyrus (B) of CBD and PSP brains, that were also positive for active caspase-6, mAbD402, and mAbD13 antibodies. CBD and PSP were selected for this analysis because they contain the greatest amount of astroglia pathology. PSP showed higher astroglia positivity for all markers, relative to CBD. Abbreviations: CBD, corticobasal degeneration; PSP, progressive supranuclear palsy. Rows represent tauopathies, controls, and a list of antibodies included in the analysis. Cells represent mean percentages of total neurons (color gradient) found positive for individual or multiple antibodies. The +/- symbols represent the presence (+, dark grey) or absence (-, light grey) of antibody signal.

### Astroglial Casp-6-cleaved tau inclusions in CBD and PSP

Tau deposits in astroglia are a prominent feature of CBD and PSP [32]. Previous studies suggest D421 tr-tau astroglial pathology [8], but it less clear if D13 and D402 tr-tau also accumulate in astroglia as well. Here, we focused on p-tau (Ser202)-positive astroglia in CBD and PSP, since these tauopathies show dominant tau lesions in this cell type. We used tissue sections that were used to quantify tau pathological changes in neurons. PSP showed a relatively strong p-tau positivity in tufted and thorned astrocytes in the MFG and ITG (Table S2; Figures 2, 6). In CBD, p-tau positivity in astrocytic plaques was also strong. In the MFG, 8.99% of p-tau-positive astrocytes also showed Casp-6 activity in PSP (3.70% in CBD). Overall, the percentages of p-tau positive astroglia with overlapping tr-tau was higher in PSP than in CBD, and D13 tr-tau was more prevalent than D402 tr-tau in both brain regions (Table S2). We failed to observe astroglia with tr-tau inclusions lacking p-tau. Our results suggest that astroglia, similar to neurons, show Casp-6-specific species of D13 and D402 tr-tau.

## Discussion

We used an established 5-plex immunofluorescence (IF) protocol [31], novel mAbs, and quantitative analyses in tissue microarray (TMA) blocks containing well-characterized postmortem brain tissue from two cortical areas of common tauopathies and healthy controls to investigate the following questions: 1) Is there evidence of aCasp-6 in non-AD tauopathies? 2) Besides D421, is accumulation of Casp-6-tr-tau fragments present in AD and other tauopathies? 3) What is the pattern of co-occurrence of N- and C-terminus tr-tau fragments, p-tau, and aCasp-6 within the same neurons? 4) Do neurons with tr-tau inclusions also show evidence of p-tau? And 5) are aCasp-6 and tr-tau inclusions also present in p-tau positive astroglia in CBD and PSP? This study offers several novel findings that inform on mechanisms and possible strategies for the diagnosis and treatment of AD and other tauopathies.

First, estimates of both Casp-6 activation and neuronal tr-tau burden were much higher in AD and, to a lesser extent, in PiD than in 4R tauopathies. In fact, evidence of Casp-6 activation was almost absent in pure 4R tauopathies (Table 2 and S2 and Figures 4, 6). aCasp-6 mediates the truncation of tau into toxic fragments prone to self-aggregation [7, 9, 19]. Thus, it is not surprising that compared to the 4R tauopathies, mean percentages of neurons with D402 and/or D13 tr-tau were 16.06- (D402; compared to PSP in MFG) to 236-fold higher in AD (D402; compared to CBD in ITG) and up to 32.67-fold higher in PiD. We previously showed that aCasp-6 inhibitors ameliorate tau pathology in iPSC-derived neurons with the frontotemporal dementia-causing V337M *MAPT* mutation [16]. Therefore, Casp inhibitors could be a promising therapy for tauopathies with 3R tau pathological forms, but they are unlikely to be useful in the treatment of 4R tauopathies.

Second, the higher tr-tau burden in AD and, to a lesser extent, in PiD was disproportionally greater than expected based on the burden of p-tau (Ser 202) inclusions. The percentage of p-tau-positive neurons in AD was 5.07x greater than that in PSP (MFG) and 3.20x greater than that in CBD (ITG). Notably, the differences between AD and PSP in the MFG and AD and PSP in the ITG were much more pronounced when comparing tr-tau burden. The percentage of tr-tau-positive neurons in AD was 16.06x greater than that in PSP (MFG) and 236x greater than that in CBD (ITG). The disproportionally higher burden of tr-tau relative to p-tau in AD compared to 4R tauopathies suggests that fluid-based biomarkers based on tr-tau may be superior to p-tau biomarkers with regards to AD diagnosis and efforts to differentiate AD from other tauopathies [33]. Developing biomarkers to detect D13 tr-tau is attractive and feasible. Although CSF lacks C-terminus tau peptides, making it challenging to detect tau fragments above residue 268 (e.g., D421) [34, 35], our multiplex IF approach enabled us to establish that an N-terminus form of tr-tau (D13) is more abundant in neurons as compared to a C-terminus form (D402). However, D13 and D402 exhibit a high degree of co-occurrence. Recent studies on fluid-based biomarkers with a N-terminal assay (NT1)—including the N-terminal mAbTau12 (amino acids 6– 13)—show that as NT1 levels increase, clinical decline in subjects with positivity for other AD biomarkers worsens. NT1 levels probably remain intact in non-AD dementia [36–38]. Supported by our quantitative neuropathological results and the success of other N-terminal truncated assays, mAbD13 may become an essential player in the arsenal of biofluid biomarkers against AD.

Third, and most intriguing, in AD, there is a population of neurons as large as the population of p-tau (Ser 202) neurons that harbors tr-tau inclusions in the absence of p-tau. P-tau aggregates are a critical feature of all tauopathies. Most clinics rely on p-tau (Ser 202) inclusions for the classification and staging of tauopathies in humans and experimental models. This is an unprecedented finding and probably highly relevant for understanding AD pathology, as neurons accumulating tr-tau may follow a pathogenic pathway independent of tau phosphorylation. Moreover, this finding suggests that there is a whole subset of neurons vulnerable to pathological tau that have been systematically excluded from investigations focusing on AD pathogenesis and the determinants of selective neuronal vulnerability because almost all studies probe p-tau alone. On the one hand, our findings may seem paradoxical because previous studies posit that tau phosphorylation precedes tau truncation in AD [8]. However, the evidence supporting this hypothesis in human brains is meager. We are only aware of a single study using postmortem samples and double-labeling to examine the co-occurrence of p-tau and tr-tau. Whereas we investigated about 2 mm^2^ of grey matter per area per case, they only examined around 0.4 mm^2^ in one area per case. This study also used TauC3 (D421), a C-terminus tr-tau, whereas N-terminus tr-tau was the most abundant form in our study. These tr-tau neurons lacking p-tau (Ser202) inclusions could be harboring inclusions that contain p-tau species which manifest in mature tangles, but our data also suggest that tangle formation may occur by distinct pathways, not necessarily including phosphorylation at Ser202. Experimental and postmortem human studies support these findings by demonstrating that tr-tau is an early event leading to filament formation in tauopathies [9, 19, 39–41]. For instance, Horowitz et al. (2004) detected N-terminal tau truncation, preceding C-terminal truncation and phosphorylation at th18 in AD [21]. Even if these authors did not have access to an antibody specific to D13 tr-tau as we have, their results support the same conclusions. Regarding tau pathogenesis, our study supports the notion that therapeutic approaches for modulating the generation of tr-tau species, including the use of aCasp-6 inhibitors or mAbD13, may be necessary to treat AD in addition to strategies aimed to modulate the accumulation of p-tau species.

Fourth, although most neurons with aCasp-6 also harbor tr-tau inclusions, the opposite is not true. In AD, almost 9/10 neurons with aCasp-6 also harbor tr-tau inclusions, while 3/4 of tr-tau inclusions show evidence of aCasp-6. This discrepancy could be attributed to a more ephemerous nature of caspase activation, as compared to tau truncation. Thus, the latter is more likely to be detected in a cross-sectional study. However, our findings in PiD suggest that the explanation for this difference is not so simple. Only about 1/3 of tr-tau neurons showed evidence of aCasp-6 in PiD. Our novel, validated monoclonal antibodies reassure that other tr-tau forms, in addition to the targeted ones, are unlikely to be detected. Thus, either the nature of Casp-6 activation is even more ephemerous in PiD than AD, which is doubtful, or, contrary to what is expected based on experimental data, other Casps, such as Casp-3, 7, or 8, may cleave tau at the D402 and D13 sites. To address this possibility, we performed immunostaining in AD tissue using antibodies against Casp-3, but found no positivity (data are not shown), in line with other studies failing to identify Casp-3 activation [14, 42, 43]. Further studies testing Casp-7 and -8 activity in tauopathies are warranted.

Fifth, D13 and D402 tr-tau was also present in p-tau-positive astroglia in PSP and, to a much lesser extent, CBD, confirming that Casp-mediated tau pathological changes extend beyond neurons in tauopathies (Figure 6, Table S2), in line with previous findings using the mAbTauC3 (D421) [8].

Our approach offers numerous strengths and includes measures to maximize analytical rigor. We generated unprecedented neoepitope mAbs against Casp-6-tr-tau at D402 (C-terminus) and D13 (N-terminus) truncation sites. We also investigated the burden of these inclusions simultaneously with p-tau and aCasp-6 burden in distinct human tauopathies. Our results highlight the importance of studies comparing different tauopathies side-by-side, as similarities and differences may inform their pathogenesis. When comparing percentages of neuronal D402 and D13 tr-tau to those of neuronal p-tau in the same disease, the differences between AD/PiD and PSP/CBD becomes even more apparent. Multiplex IF methods have been limited by difficulties in eluting any antibody, including tau antibodies, while maintaining tissue integrity to allow for multiple staining cycles in the same histological slide. Here, we applied a pipeline recently developed in-house that provides excellent elution of tau antibodies verified by rigorous quality control steps [31]. Our methodology enabled us to examine three tau antibodies raised against the same species simultaneously. Additionally, the simultaneous use of several antibodies in the same tissue section increases confidence in our results. To avoid potential biases stemming from immunostaining batch variation, we produced TMAs containing multiple brain specimens in a single paraffin block. This approach also minimizes the chances of false-negative results. For instance, all our tissue samples, except for AGD which is not expected to show p-tau positivity in the MFG [44, 45], showed a relatively strong p-tau positivity, and CBD almost lacked evidence of Casp-6 activation when compared to the other punches in the same TMA (Table 2; Figures 4, 5). TMAs are a commonly used approach in cancer research but are still a developing approach in research on neurodegenerative diseases. Finally, we quantified neurons and pathological inclusions in two separate sets of slides of the same case cases, with similar results. Nevertheless, our approach is not free of limitations, many inherent to postmortem studies involving the human brain. Such limitations include the cross-sectional nature of the specimens and the relatively small number of cases analyzed. Obtaining well-characterized postmortem human brain tissue from healthy controls and rare tauopathies, especially those from cases lacking neuropathological comorbidities, is challenging. Despite our relatively small cohort size, we counted over 40,000 individual neurons in MFG and ITG combined.

In summary, this study demonstrates a strong association between aCasp-6 and tr-tau, suggesting that Casp-6-cleavage of tau in neurons is likely a feature of tauopathies with 3R, rather than 4R, tau. In AD, sizeable percentages of neurons positive for tr-tau lacked evidence of p-tau. Considering the progressive nature of AD and other tauopathies, early modulation of Casp-6 activation and Casp cleavage of tau could have a significant therapeutic value against tau aggregation and neuronal death. This study further supports the potential of biofluid biomarkers for tr-tau forms to diagnose AD and differentiate AD from other tauopathies at the single patient level. Future studies in human and clinically-relevant models of tau pathology, such as iPSCs from patients with MAPT mutations [46], are crucial for a better understanding of caspase-mediated pathways that lead to tau pathology in tauopathies.

## Data Availability

The data that support the findings of this study are available from the corresponding author upon reasonable request.

## Abbreviations

3R: 3-repeat
4R: 4-repeat
aCasp-6: Active caspase-6
AD: Alzheimer’s disease
AGD: Argyrophilic grain disease
CBD: Corticobasal degeneration
IF: Immunofluorescence
ITG: Inferior temporal gyrus
MFG: Middle frontal gyrus
mAb: Monoclonal antibody
NFT: Neurofibrillary tangle
p-tau: Hyperphosphorylated tau
PHF: Paired helical filaments
PiD: Pick’s disease
PSP: Progressive supranuclear palsy
PTM: Post-translational modifications
tr-tau: Truncated tau
TMA: Tissue microarrays

## Author Contributions

P.T., A.M.H.P., and L.T.G. designed the study; P.T. and A.M.H.P., performed the experiments, data acquisition and analyses. B.C.,T.Y., S.K., R.N., and M.R.A. generated the neoepitope caspase-cleaved tau antibodies. S.L, A.J.E, and R.A.E contributed to immunohistochemistry and C.P. with data analyses and graphic design. S.S, W.W.S., B.L.M, and L.T.G provided the human brain specimens, P.T. and A.M.H.P. wrote the manuscript, S.S, W.W.S., B.L.M, M.R.A., and L.T.G. revised the manuscript, L.T.G. supervised the study and approved the submitted version.

## Acknowledgements

We thank the patients and their families for their invaluable contribution to brain ageing neurodegenerative disease research. We thank Dr Andrea LeBlanc for helpful discussions on caspase biochemistry and Dr Peter Davies (Albert Einstein College of Medicine, New York, NY) for generously providing tau antibodies. This study was supported by the National Institutes of Health K01AG053433 (P.T), K24AG053435, R56AG057528 and U54 NS100717 (L.T.G), K08AG052648 (S.S), P30AG062422 and P01AG019724 (B.L.M), UCSF RAP Pilot Award program (P.T), UCSF RAP Team Science Grant (L.T.G., M.R.A.), Alzheimer’s Association AARG-16-441514 (L.T.G., M.R.A.), Rainwater Charitable Foundation (M.R.A.), and a Catalyst award from ShangPharma Innovation (M.R.A., T.Y., S.K, R.N.).

## Ethical approval

Use of human brain tissue has received ethical approval by the University of California San Francisco Institutional Review Board.

## References

1. Spillantini MG, Goedert M (2013) Tau pathology and neurodegeneration. Lancet Neurol 12:609–622

2. Rösler TW, Tayaranian Marvian A, Brendel M, et al (2019) Four-repeat tauopathies. Prog Neurobiol 180:101644

3. Cairns NJ, Bigio EH, Mackenzie IRA, et al (2007) Neuropathologic diagnostic and nosologic criteria for frontotemporal lobar degeneration: consensus of the Consortium for Frontotemporal Lobar Degeneration. Acta Neuropathol (Berl) 114:5–22

4. Morris M, Maeda S, Vossel K, Mucke L (2011) The Many Faces of Tau. Neuron 70:410–426

5. Tracy T, Claiborn KC, Gan L (2019) Regulation of Tau Homeostasis and Toxicity by Acetylation. Adv Exp Med Biol 1184:47–55

6. Wesseling H, Mair W, Kumar M, et al (2020) Tau PTM Profiles Identify Patient Heterogeneity and Stages of Alzheimer’s Disease. Cell 183:1699–1713.e13

7. de Calignon A, Fox LM, Pitstick R, Carlson GA, Bacskai BJ, Spires-Jones TL, Hyman BT (2010) Caspase activation precedes and leads to tangles. Nature 464:1201–1204

8. Ferrer I, Lopez-Gonzalez I, Carmona M, Arregui L, Dalfo E, Torrejon-Escribano B, Diehl R, Kovacs GG (2014) Glial and Neuronal Tau Pathology in Tauopathies: Characterization of Disease-Specific Phenotypes and Tau Pathology Progression. J Neuropathol Exp Neurol 73:17

9. Gamblin TC, Chen F, Zambrano A, et al (2003) Caspase cleavage of tau: linking amyloid and neurofibrillary tangles in Alzheimer’s disease. Proc Natl Acad Sci U S A 100:10032–10037

10. Zhao Y, Tseng I-C, Heyser CJ, et al (2015) Appoptosin-Mediated Caspase Cleavage of Tau Contributes to Progressive Supranuclear Palsy Pathogenesis. Neuron 87:963–975

11. Gray DC, Mahrus S, Wells JA (2010) Activation of specific apoptotic caspases with an engineered small-molecule-activated protease. Cell 142:637–646

12. Hyman BT, Yuan J (2012) Apoptotic and non-apoptotic roles of caspases in neuronal physiology and pathophysiology. Nat Rev Neurosci 13:395–406

13. Albrecht S, Bourdeau M, Bennett D, Mufson EJ, Bhattacharjee M, LeBlanc AC (2007) Activation of caspase-6 in aging and mild cognitive impairment. Am J Pathol 170:1200–1209

14. Guo H, Albrecht S, Bourdeau M, Petzke T, Bergeron C, LeBlanc AC (2004) Active caspase-6 and caspase-6-cleaved tau in neuropil threads, neuritic plaques, and neurofibrillary tangles of Alzheimer’s disease. Am J Pathol 165:523–531

15. Nikolaev A, McLaughlin T, O’Leary DDM, Tessier-Lavigne M (2009) APP binds DR6 to trigger axon pruning and neuron death via distinct caspases. Nature 457:981–989

16. Theofilas P, Ehrenberg AJ, Nguy A, et al (2018) Probing the correlation of neuronal loss, neurofibrillary tangles, and cell death markers across the Alzheimer’s disease Braak stages: a quantitative study in humans. Neurobiol Aging 61:1–12

17. Dagbay KB, Hardy JA (2017) Multiple proteolytic events in caspase-6 self-activation impact conformations of discrete structural regions. Proc Natl Acad Sci 114:E7977–E7986

18. Graham RK, Ehrnhoefer DE, Hayden MR (2011) Caspase-6 and neurodegeneration. Trends Neurosci 34:646–656

19. Rissman RA, Poon WW, Blurton-Jones M, Oddo S, Torp R, Vitek MP, LaFerla FM, Rohn TT, Cotman CW (2004) Caspase-cleavage of tau is an early event in Alzheimer disease tangle pathology. J Clin Invest 114:121–130

20. Foveau B, Albrecht S, Bennett DA, Correa JA, LeBlanc AC (2016) Increased Caspase-6 activity in the human anterior olfactory nuclei of the olfactory bulb is associated with cognitive impairment. Acta Neuropathol Commun 4:127

21. Horowitz PM, Patterson KR, Guillozet-Bongaarts AL, Reynolds MR, Carroll CA, Weintraub ST, Bennett DA, Cryns VL, Berry RW, Binder LI (2004) Early N-terminal changes and caspase-6 cleavage of tau in Alzheimer’s disease. J Neurosci Off J Soc Neurosci 24:7895–7902

22. Ramcharitar J, Albrecht S, Afonso VM, Kaushal V, Bennett DA, Leblanc AC (2013) Cerebrospinal fluid tau cleaved by caspase-6 reflects brain levels and cognition in aging and Alzheimer disease. J Neuropathol Exp Neurol 72:824–832

23. Eser RA, Ehrenberg AJ, Petersen C, et al (2018) Selective Vulnerability of Brainstem Nuclei in Distinct Tauopathies: A Postmortem Study. J Neuropathol Exp Neurol 77:149–161

24. Montine TJ, Phelps CH, Beach TG, et al (2012) National Institute on Aging-Alzheimer’s Association guidelines for the neuropathologic assessment of Alzheimer’s disease: a practical approach. Acta Neuropathol (Berl) 123:1–11

25. Mackenzie IRA, Neumann M, Bigio EH, et al (2010) Nomenclature and nosology for neuropathologic subtypes of frontotemporal lobar degeneration: an update. Acta Neuropathol (Berl) 119:1–4

26. Theofilas P, Wang C, Butler D, et al (2021) Caspase inhibition mitigates tau cleavage and neurotoxicity in iPSC-induced neurons with the V337M MAPT mutation. bioRxiv 2021.01.08.425912

27. Espinoza M, de Silva R, Dickson DW, Davies P (2008) Differential incorporation of tau isoforms in Alzheimer’s disease. J Alzheimers Dis JAD 14:1–16

28. Goedert M, Jakes R, Crowther RA, Six J, Lübke U, Vandermeeren M, Cras P, Trojanowski JQ, Lee VM (1993) The abnormal phosphorylation of tau protein at Ser-202 in Alzheimer disease recapitulates phosphorylation during development. Proc Natl Acad Sci U S A 90:5066–5070

29. Jicha GA, Weaver C, Lane E, Vianna C, Kress Y, Rockwood J, Davies P (1999) cAMP-dependent protein kinase phosphorylations on tau in Alzheimer’s disease. J Neurosci Off J Soc Neurosci 19:7486–7494

30. Zhou X-W, Li X, Bjorkdahl C, Sjogren MJ, Alafuzoff I, Soininen H, Grundke-Iqbal I, Iqbal K, Winblad B, Pei J-J (2006) Assessments of the accumulation severities of amyloid β-protein and hyperphosphorylated tau in the medial temporal cortex of control and Alzheimer’s brains. Neurobiol Dis 22:657–668

31. Ehrenberg AJ, Morales DO, Piergies AMH, Li SH, Tejedor JS, Mladinov M, Mulder J, Grinberg LT (2020) A manual multiplex immunofluorescence method for investigating neurodegenerative diseases. J Neurosci Methods 108708

32. Coughlin DG, Dickson DW, Josephs KA, Litvan I (2021) Progressive Supranuclear Palsy and Corticobasal Degeneration. Adv Exp Med Biol 1281:151–176

33. Karikari TK, Pascoal TA, Ashton NJ, et al (2020) Blood phosphorylated tau 181 as a biomarker for Alzheimer’s disease: a diagnostic performance and prediction modelling study using data from four prospective cohorts. Lancet Neurol 19:422–433

34. Cicognola C, Brinkmalm G, Wahlgren J, et al (2019) Novel tau fragments in cerebrospinal fluid: relation to tangle pathology and cognitive decline in Alzheimer’s disease. Acta Neuropathol (Berl) 137:279–296

35. Sato C, Barthélemy NR, Mawuenyega KG, et al (2018) Tau kinetics in neurons and the human central nervous system. Neuron 97:1284–1298.e7

36. Chen Z, Mengel D, Keshavan A, et al (2019) Learnings about the complexity of extracellular tau aid development of a blood-based screen for Alzheimer’s disease. Alzheimers Dement J Alzheimers Assoc 15:487–496

37. Chhatwal JP, Schultz AP, Dang Y, Ostaszewski B, Liu L, Yang H-S, Johnson KA, Sperling RA, Selkoe DJ (2020) Plasma N-terminal tau fragment levels predict future cognitive decline and neurodegeneration in healthy elderly individuals. Nat Commun 11:6024

38. Mengel D, Janelidze S, Glynn RJ, Liu W, Hansson O, Walsh DM (2020) Plasma NT1 Tau is a Specific and Early Marker of Alzheimer’s Disease. Ann Neurol 88:878–892

39. Guillozet-Bongaarts AL, Glajch KE, Libson EG, Cahill ME, Bigio E, Berry RW, Binder LI (2007) Phosphorylation and cleavage of tau in non-AD tauopathies. Acta Neuropathol (Berl) 113:513–520

40. Canu N, Dus L, Barbato C, Ciotti MT, Brancolini C, Rinaldi AM, Novak M, Cattaneo A, Bradbury A, Calissano P (1998) Tau cleavage and dephosphorylation in cerebellar granule neurons undergoing apoptosis. J Neurosci Off J Soc Neurosci 18:7061–7074

41. Delobel P, Lavenir I, Fraser G, Ingram E, Holzer M, Ghetti B, Spillantini MG, Crowther RA, Goedert M (2008) Analysis of Tau Phosphorylation and Truncation in a Mouse Model of Human Tauopathy. Am J Pathol 172:123–131

42. LeBlanc A, Liu H, Goodyer C, Bergeron C, Hammond J (1999) Caspase-6 role in apoptosis of human neurons, amyloidogenesis, and Alzheimer’s disease. J Biol Chem 274:23426–23436

43. Selznick LA, Holtzman DM, Han BH, Gökden M, Srinivasan AN, Johnson EM, Roth KA (1999) In situ immunodetection of neuronal caspase-3 activation in Alzheimer disease. J Neuropathol Exp Neurol 58:1020– 1026

44. Grinberg LT, Heinsen H (2009) Argyrophilic grain disease: an update about a frequent cause of dementia. Dement Neuropsychol 3:2–7

45. Kovacs GG (2015) Invited review: Neuropathology of tauopathies: principles and practice. Neuropathol Appl Neurobiol 41:3–23

46. Karch CM, Kao AW, Karydas A, et al (2019) A Comprehensive Resource for Induced Pluripotent Stem Cells from Patients with Primary Tauopathies. Stem Cell Rep 13:939–955

