## Supplementary material for "Caspase-6-cleaved tau is relevant in Alzheimer’s disease but not in other tauopathies: diagnostic and therapeutic implications"

**Lea T. Grinberg, MD, Ph.D**; John Douglas French Alzheimer's Foundation Endowed Professor;

**Supplemental Experimental Procedures**

**A. Development of caspase-6 cleaved tau neoepitope antibodies**

**Analysis of antibody specificity**

To evaluate the specificity of mAbD402 and mAbD13 in detecting tr-tau, we performed antigen competition assays using immunohistochemistry (IHC). We used the peptides as immunogens for antibody generation on adjacent brain sections. Two IHC experiments were run in duplicates. One slide was pre-incubated with mAbD402 (or mAbD13) immunogenic peptides in the primary antibody step, and the other two slides were incubated with untreated mAbD402 or mAbD13. All other experimental parameters remained constant. In each reaction, we also included slides in which the primary antibody was replaced by saline buffer as a negative control. We observed immunoperoxidase signal only in the slides incubated with the primary antibody in the cognate immunogenic peptide's absence [1].

**B. Tissue processing and multiplex immunofluorescence staining**

**Antibody labeling**

Tissue sections were first incubated with NeuN (1:300) at room temperature for one hour, followed by goat anti-guinea pig HRP (1:200, Advansta). NeuN signal was developed using Ventana's Discovery DCC kit.

An antibody denature step was used to remove excess unbound secondary HRPs by incubating sections in 95°C BenchMark Ultra CC2 solution for eight minutes. Next, sections were incubated with mAbD13 (1:150) overnight. The Discovery anti-mouse HQ, anti-HQ HRP, and FAM kit were used to develop the mAbD13 signal. Sections were then stripped of mAbD13 and incubated with CP13 (1:2000) at room temperature for three hours. CP13 signal was developed using the Discovery omnimap anti-mouse HRP and Cy5 kit. To avoid cross-reaction between primary antibodies from the same species, an antibody stripping step (using BenchMark Ultra CC2 solution at high heat and the Discovery antibody denaturing reagent) was performed following tyramide-signal amplification (TSA) development of mAbD13 and CP13. Finally, sections were incubated with the remaining primary antibodies—a cocktail of active caspase-6 (1:800) and mAbD402 (1:700)—at room temperature for six hours. The active caspase-6 signal was developed with Discovery omnimap anti-rabbit HRP and Rhod 6G kit. mAbD402 signal was developed with biotinylated anti-mouse (1:200, Vector Laboratory) with Streptavidin Alexa Fluor 790 (1:200, Thermo Fisher). To reduce autofluorescence, sections were incubated in Sudan Black B (Sigma), and coverslipped with prolong antifade mounting media (Invitrogen).

**C. Quantitative analysis of positive markers**

We manually quantified neurons (NeuN-positive) in all tauopathies using FIJI's built-in counting tool [2]. We followed the same approach for quantifying pathological tau inclusions in astroglia based on p-tau (mAbCP13) positivity in PSP and CBD. These diseases show the most pronounced astroglial pathology in our tauopathy cohort. Next, we counted the number of cells positive for each marker on stacked, single-channel images. FIJI's counting tool tracks the number of counters placed within each image, and different counter types can be used to track cells that the user classifies as positive for various markers. Based on an oversampling-resampling analysis [3, 4], we counted approximately 1000 cells per section from every fourth 500 x 500 μm box containing the gray matter randomly sampled from a grid overlaying the images. In sections containing less grey matter, we increased the number of boxes counted to reach 1000 cells. Next, we used an in-house developed Python script to automatically determine marker co-occurrence based on counter data extracted from FIJI files (Figure 3). The script reads in counter coordinates, which are grouped by the marker. It stores them in marker-specific dictionaries (i.e., coordinates for counters placed on mAbD402-positive cells are stored in one dictionary, and coordinates for markers placed on mAbD13-positive cells are stored in another dictionary). The script then determines the number of co-occurring markers by searching between the dictionaries for coordinates within a two-pixel radius. We quantified two subsets of TMAs (same TMA and areas, but different staining batches) to increase the rigor, and the results were similar.

**References**

1.	Theofilas P, Wang C, Butler D, et al (2021) Caspase inhibition mitigates tau cleavage and neurotoxicity in iPSC-induced neurons with the V337M MAPT mutation. bioRxiv 2021.01.08.425912

2.	Schindelin J, Arganda-Carreras I, Frise E, et al (2012) Fiji: an open-source platform for biological-image analysis. Nat Methods 9:676–682

3.	Slomianka L, West MJ (2005) Estimators of the precision of stereological estimates: an example based on the CA1 pyramidal cell layer of rats. Neuroscience 136:757–767

4.	Theofilas P, Polichiso L, Wang X, et al (2014) A novel approach for integrative studies on neurodegenerative diseases in human brains. J Neurosci Methods 226:171–183

| **Primary antibody** | **Host** | **Dilution** | **Provider** |
| --- | --- | --- | --- |
| **Active caspase-6 (Asp179)** | Polyclonal rabbit | 1:500 | Aviva System Biology (Cat Nr: OAAF05316) |
| **Phosphorylated tau (CP13; Ser 202)** | Monoclonal mouse | 1:400 | Gift from Peter Davies, NY |
| **Caspase-6-cleaved tau (D13; N-terminus)** | Monoclonal mouse | 1:200 | Developed in house |
| **Caspase-6-cleaved tau (D402; C-terminus)** | Monoclonal mouse | 1:200 | Developed in house |
| **NeuN** | Polyclonal guinea pig | 1:300 | Synaptic System (Cat Nr: 266 004) |

**Supporting Table S1.** List of antibodies used for multiplex immunofluorescent staining.

|  | **Middle frontal gyrus Inferior temporal gyrus** | | | | |
| --- | --- | --- | --- | --- | --- |
| **Variable** | **CBD** | **PSP** |  | **CBD** | **PSP** |
| **n** | 3 | 3 |  | 3 | 3 |
| **Number of astroglia**  **(p-tau)** | 36.33  (32.32) | 49.67  (28.75) |  | 17.00  (5.20) | 6.67  (3.21) |
| **Astroglial aCasp-6 (%)** | 3.70  (6.42) | 8.99  (7.41) |  | 0.00  (0.00) | 8.33  (14.43) |
| **Astroglial D402 tr-tau (%)** | 0.00  (0.00) | 6.14  (5.65) |  | 0.00  (0.00) | 12.50  (21.65) |
| **Astroglial D13 tr-tau (%)** | 5.09  (5.61) | 14.72  (12.75) |  | 0.00  (0.00) | 19.44  (17.35) |
| **aCasp-6 + D402 tr-tau (%)** | 0.00  (0.00) | 2.44  (4.22) |  | 0.00  (0.00) | 8.33  (14.43) |
| **aCasp-6 + D13** **tr-tau (%)** | 3.70  (6.42) | 5.30  (6.25) |  | 0.00  (0.00) | 8.33  (14.43) |
| **D402 tr-tau + D13 tr-tau (%)** | 0.00  (0.00) | 6.14  (5.65) |  | 0.00  (0.00) | 8.33  (14.43) |
| **aCasp-6 + D402 tr-tau + D13** **tr-tau (%)** | 0.00  (0.00) | 2.44  (4.22) |  | 0.00  (0.00) | 8.33  (14.43) |

**Supporting Table S2. Mean (SD) percent of p-tau-positive astroglia also positive for the individual markers active caspase-6, D402 truncated tau, D13 truncated tau, individually and by each combination of co-occurring markers.** PSP cases showed higher percentages of astroglia positive for active caspase-6, mAbD402, and mAbD13 in both brain regions analyzed compared to CBD cases. Numbers for individual markers include astroglia with more than one marker Abbreviations: aCasp-6, active caspase-6; CBD, corticobasal degeneration; PSP, progressive supranuclear palsy; p-tau, phosphorylated tau; mAb, monoclonal antibody.
